# Trans-border transfer of human biological materials in collaborative biobanking research: Perceptions and experiences of researchers in Uganda

**DOI:** 10.1101/2022.04.01.22273073

**Authors:** Erisa Sabakaki Mwaka, Ian Guyton Munabi

**Author notes:** Emails: EMS IGM. Both EMS and IGM participated equally as first authors. Corresponding Author: Erisa Sabakaki Mwaka.

## Abstract

**Introduction:** The study aimed to explore researchers’ perceptions and experiences on the transfer of human biological materials in international collaborative research.

**Methods:** This was a descriptive convergent parallel survey that randomly recruited 187 researchers involved in biobanking and/or genetics research. Data were collected using a self-administered tool that had both open and closed ended questions.

**Results:** Majority of respondents were male scientists (53.5%) with a mean research experience of 12.2 (SD 6.75) years. About 89% had ever worked in international collaborative research and 42% had ever participated in material transfer agreement (MTA) development. There were several areas of agreement in regard to the rights of local researchers and institution in collaborative biobanking research, and what details should be included in material transfer agreements. There was overwhelming support for collaborative partnership in biobanking research with 90.4% of researchers agreeing that local scientists should be involved in decisions making regarding the future use of samples. A majority (85.6%) opined that there should be fair sharing of research benefits with local researchers and populations or country from which the human biological materials were taken. Researchers felt that most MTAs tend to favour international collaborators. Several trust issues in the MTA development and implementation process were also highlighted and these included: lack of transparency and dishonesty of receiving scientists; lack of mechanisms to monitor the use of exported samples; ownership and intellectual property disputes; exploitation and inequitable sharing of research benefits; and authorship challenges. Several researchers seemed not to be conversant with the guidance provided by the Ugandan national ethics guidelines on the cross-border exchange of human biological specimens.

**Conclusion:** Local researchers had a positive attitude towards the export of human biological materials in collaborative research. However, there were several governance and trust concerns. There is a need for collaborative partnership in biobanking research.

## Introduction

For decades, large volumes of human biological materials (HBM) and data have been exported from developing to developed countries for storage in biorepositories and future unknown research [1]. This practice has raised pertinent and legitimate questions, and concerns for scientists, research participants and funders [2, 3]. Since inception of the Human Heredity and Health in Africa (H3Africa) initiative [4], there has been an exponential growth in biobanking and genetics research on the African continent and vast quantities of HBM and data have been collected. H3Africa is a National Institutes of Health and Wellcome Trust funded initiative that is spearheading biobanking and genomics research in Africa for Africa and is providing new opportunities to African scientists to lead research on the genetic and environmental contributors to health and common disease issues of importance to Africa through the use of genomics and other cutting-edge approaches [1]. Currently there are six H3Africa research projects in Uganda including the Integrated Biorepository of H3Africa Uganda (IBRH3AU) that was established in 2010. The biorepository is custodian to than 300,000 HBM from more than 100,000 participants [5].

The need to share samples across national borders is increasingly becoming important because of widespread north-south collaboration and commercial activities [6]. As a result, large quantities of HBM and data have been shipped from low resource-to developed countries for storage because of lack of biobanks in Africa [1]. There is empirical evidence that samples may be crossing borders without material transfer agreements (MTA) and/or export permits in African collaborative research [7], and this has been equated to ‘trafficking of samples.’ The fate of these HBM is unknown because regulatory authorities/bodies from most resource limited countries have no mechanisms for monitoring HBM once they cross their borders. This situation is further exacerbated by the lack of comprehensive ethico-legal frameworks and other weaknesses in biobank governance [8]. Absence of appropriate regulatory frameworks exposes HBM to abuse and exploitation. For example, during the 2014-2016 Ebola virus disease epidemic in West Africa, HBM were exported from Sierra Leone and Liberia for storage in other countries without valid informed consent. The custodians of those particular samples have also persistently resisted any form of regulatory oversight or access to those samples by researchers and governments of the countries from which they were collected [9, 10]. Such incidents could cause misconception and pose a significant risk to public trust in research [11, 12] as well as biobanking [13–15]. Further, over the years, several biobank governance misconducts by researchers from developed countries have been highlighted in literature. There have been accusations of contravention of national guidelines [16], cultural indifference [17], fraud and theft of intellectual property [18], commercialization of HBM and data without informed consent [19, 20], exploitation of participants [21] inequitable benefit sharing [22–24], exclusion of local investigators from publication authorship [25, 26], loss of revenue to uncontrolled export of HBM [27] and allegations of viewing African institutions and their researchers as specimen collecting centres and collection technicians respectively [28]. In the initial years of the H3Africa initiative it was realized that most countries in sub-Saharan Africa had no comprehensive national guidance for biobanking and genetics research [8]. The H3Africa Consortium has tried to tackle some of these gaps by infrastructural and human capacity strengthening, and by developing several guidance documents on ethics, governance and resource sharing to foster best ethical practices within the consortium [3,29–35].

Most research in Uganda is foreign funded and this presents unique challenges in international collaborations [33] because of the differing national positions that make implementation of national laws consistently difficult [34]. Research in Uganda is guided by the National Guidelines for Research involving Humans as Research Participants [35] and the National Research Biobanking Guidelines [36] that provide guidance on the acquisition, storage, and the exchange/transfer of HBM [35]. However, these guidelines are not enshrined in law, are brief and lack detail as compared to other countries such as South Africa [37]. There is a need to explore sources of contention to develop a consistent and equitable framework for collaborative biobank research in low and middle-income countries (LMIC). The World Health Organization recommends increased involvement and recognition of contribution of scientists from countries of origin in research and specimen utilization, and fair representation in scientific publications [38]. Researchers and policy makers opine that intellectual property rights for research on exported HBM should be shared with local scientists and communities [24]. Local principal investigators also would like to be consulted whenever any samples exported from their repositories are used for new research [24]. With this background, this study aimed to explore researchers’ perceptions and experiences of transferring human biological materials in international collaborative research.

## Methods

This was a descriptive convergent mixed methods design study [39], study looking at Ugandan researcher’s perceptions and experiences of sharing of HBM in collaborative research. The quantitative component of the data was obtained from a set of closed ended items looking at various aspects of the sample sharing experience, while the qualitative data came from an open-ended item.

### Study population

The study made use of the national database of researchers held by the national research regulator, the Uganda National Council for Science and Technology (UNCST) that had 2,354 registered research protocols on its records at the start of the study (November 11 2016). From this data set of previously submitted protocols by the national research regulator (UNCST), a manual search was made for protocols proposing either to store HBM for future use in research (biobanking) and/or conduct genomic/ genetics related analyses were identified for inclusion in the study. The investigators associated with these protocols were compiled into a list of potential participants. Identified investigators could only appear once on the list even if they had submitted more than one biobanking related research to UNCST. A set of computer generated random numbers was then used to select which researchers to contact for inclusion in the study from this list.

### Sample size

The target sample size for the study was obtained assuming that 50% of the research protocols were focused on health-related research that typically involves human biological samples or specimens that may later be used in genomic or genetics related research, for a design effect of 1.2 due to the multiple institutions represented in the pool or protocols, confidence limits of 5% and power of 80% from a potential pool of 2,354 researcher protocols. This gave a final sample size of 185 researchers to which was included a 15 percent allowance for loss and or non-response to give a final target rounded up sample size of 213 participants. This sample size was obtained using the online calculator for sample size based on proportions available on www.openepi.com website.

### Instruments

The survey tool to be used will be adapted from the International survey of scientist and policy makers’ attitudes toward research on stored human biological materials [24]. It explores consent options for the storage and future use of samples; requirements for re-contacting participants; and data and sample sharing in international collaborative research. The tool had one open ended item asking respondents to provide any comments they might have wanted to share about their experience with using MTAs.

### Study procedure

A physical search was made of the regulators protocol archive for all health-related research protocols ranging from 2012 year to 2017. The identified health related research protocols were further screened to identify those that included biobanking or planned to use samples for further genomic or genetic analysis. These investigators were randomly selected for inclusion in the study using computer generated numbers. The selected researchers were contacted and invited to participate in the study. The researchers who accepted to participate in the study were given the study self-administered questionnaire after providing written informed consent. Participants were given up to three reminders to return the duly completed study questionnaires by one of the study research assistants. Those that had not completed the questionnaire after the third reminder were considered as non-responsive or having withdrawn their participation. Participants who returned the completed study questionnaire were given a token of appreciation which also marked the end of their participation in the study.

### Data analysis

The completed questionnaires were digitized using epidata 3.1 by a qualified and trained data entrant. The digitized study records file was saved as a RDS file for further data cleaning and analysis using R version 4.1.2 [40]. The open-ended items were exported separately as text files for qualitative analysis in R version 3.5.3 [41], using the RQDA [42] and tidy text [43] packages. Quantitative data analysis: Descriptive statistics for each of the variables in the data set were generated. Subsequent analysis focused on determining the association between the different variables and the respondents reported involvement in sample collection for research using odds ratios. The results of the quantitative analyses were summarized as tables capturing different descriptive statistics and the odds ratios. The level of significance for all the quantitative statistical tests was set at 0.05. Qualitative data analysis: The responses of participants to the open-ended questions were edited to correct spelling mistakes then subjected to content analysis to identify respondents’ sentiments towards their experiences with the MTA according to gender, after removal of all stop words. The sentiment analysis made use of the “bing” database found in the tidy text r package [43, 44]. Following the sentiment analysis, a word cloud was generated as a summary of the top 200 most frequently used words in the respondent’s feedback. This was followed by a thematic analysis [45], of the respondents’ responses involving multiple readings of the text by both authors initially grouped into codes and themes. During these rounds of reading the authors kept a record of their thoughts and impressions on reading the text using memos that informed the revision and reorganizing of the generated information leading to the final set of codes and themes included in the results section. The information from the memos, capturing the authors impressions, views and opinions about the data, was used in the creation of the descriptions that accompany each of the generated codes and themes. each description included a summary of the number of actual respondents’ comments and an example of a quote under each of the codes. The mixing of the two sets of data from the qualitative and quantitative arms of the study was done during the discussion to provide a rich description of the study observations as part of a convergent mixed methods design [39]. The R code and corresponding data for all the analysis was made available online [46].

### Ethical considerations

Ethics approval was obtained from the Makerere University School of Biomedical Sciences Research Ethics Committee (SBS-517) and Uganda National Council for Science and Technology (SS 4190). Informed consent was obtained from all respondents prior to enrolment in the study. Participants were assured of confidentiality; no participant personal identifier marks or names were collected. Each participant received a token compensation of 15 USD for the time spent participating in the study.

## Results

As shown in the participant flow diagram (see figure 1), 3,298 protocols were registered by UNCST for the period January 2012 to December 2017. Only 243 of these protocols either involved biobanking or genomic/genetics research. Of these 213 were selected following randomization and one investigators was selected per protocol for inclusion in the study. At the end of the study period only 187 researchers had provided their informed consent and returned the completed study questionnaires.

**Figure 1:**
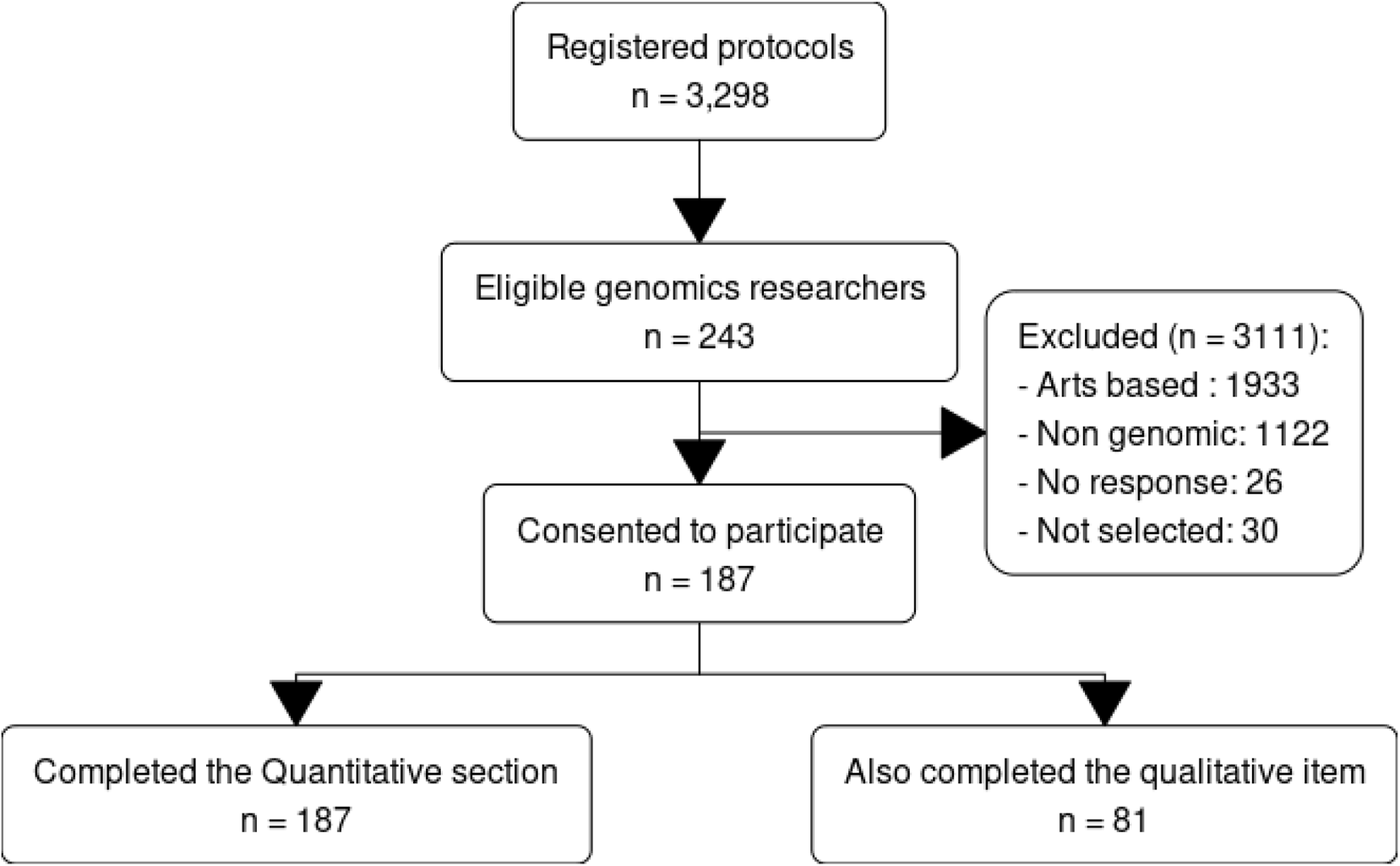
Participant flow diagram

The majority of the respondents were males (100/187) for this survey. The average age of the participants was 42.33, (SD=10.61) years as shown in Table 1. The respondents described themselves as scientists (85%) and others identified themselves as coming from academia (34.2%), clinical medicine (43.9%), health policy (3.7%), and affiliated to research ethics committees (REC) (9.6%). On average participants had been involved in research for 12.16, (SD=6.75) years. The reported highest level of education also varied with the majority having attained masters level training (68.4%) followed by PhD (26.7%) and the rest were bachelor’s degree holders (9%). It is important to note that some researchers reported multiple occupational affiliations.

**Table 1:** Summary statistics of the study population

| values | Observations<br>(N=187) |
| --- | --- |
| **Gender** |  |
| Male | 100 (53.5%) |
| Female | 87 (46.5%) |
| **Age** |  |
| Mean (SD) | 42.3 (10.6) |
| Median [Min, Max] | 41.0 [26.0, 99.0] |
| **Years Worked in research field** |  |
| Mean (SD) | 12.2 (6.75) |
| Median [Min, Max] | 12.0 [1.00, 34.0] |
| **Conducting research on stored human biological samples** |  |
| Yes | 129 (69.0%) |
| No | 55 (29.4%) |
| **Involved in the collection of human biological samples for use in future research** |  |
| Yes | 162 (86.6%) |
| No | 25 (13.4%) |
| **Have ever worked in collaborative partnerships with people from other countries** |  |
| Yes | 166 (88.8%) |
| No | 21 (11.2%) |
| **12. Have you collaborated with people in:** |  |
| Only developing countries | 4 (2.1%) |
| Only developed countries | 24 (12.8%) |
| Both developing and developed co | 139 (74.3%) |
| **Clinical medicine** |  |
| Yes | 82 (43.9%) |
| **Scientific research** |  |
| Yes | 159 (85.0%) |
| **REC** |  |
| Yes | 18 (9.6%) |
| **Academia** |  |
| Yes | 64 (34.2%) |
| **Health policy** |  |
| Yes | 7 (3.7%) |

### Involvement in biobanking research

From table 2 it is also important to note that the participants who were involved in collecting of HBM for future research were also involved in conducting research on stored HBM. This remained significant after controlling for the participants gender, having themselves donated samples, and them currently conducting research on stored HBM (Adjusted Odds ratio = 9.09(3.25-28.87, *p* < 0.001)). Regarding the participants’ role in the use of MTA’s, 42 % of the respondents said they had been involved in the development of the MTA’s. There were 9 % who indicated that they were the ones receiving the samples. On the other hand, 57 % said they were the ones involved in the transfer of samples. Finally, 33 % of the study participants said that they had never been involved in the use of MTA’s. Table 3, provides comparisons of the study participants perceptions of the MTA handling process based on their reported involvement in sample collection research. As shown in table 1, 86.6% of the respondents said they were collecting biological samples for use in future research. In table 2 we note that participants who were involved in the collection of human biological samples were three (3) times more likely to be involved in collaborative partnerships compared to those that said no to this question. This was not significant after controlling for the participants gender, having themselves donated samples, and them currently conducting research on stored samples (Adjusted Odds ratio = 2.71(0.81-8.77, p = 0.096)).

**Table 2:** Comparisons based on reported involvement in sample collection research

| Dependent: Involved in the collection of human biological samples for use in future research |  | Yes | No | OR (univariable) | OR (multivariable) |
| --- | --- | --- | --- | --- | --- |
| Gender | Male | 92 (92.0) | 8 (8.0) | - | - |
| | Female | 70 (80.5) | 17 (19.5) | 2.79 (1.17-7.19, $p=0.025$) | 1.97 (0.73-5.64, $p=0.188$) |
| | No | 36 (65.5) | 19 (34.5) | 10.82 (4.23-31.59, $p<0.001$) | 9.09 (3.25-28.87, $p<0.001$) |

| Dependent: Involved in the collection<br>of human biological samples for use in<br>future research |  | Yes | No | OR (univariable) | OR<br>(multivariable) |
| --- | --- | --- | --- | --- | --- |
| Have ever donated biological samples<br>for research purposes | Yes | 41<br>(87.2) | 6<br>(12.8) | - | - |
|  | No | 118<br>(86.1) | 19<br>(13.9) | 1.10 (0.43-3.19,<br>p=0.849) | 0.51 (0.16-1.70,<br>p=0.260) |
| Have ever worked in collaborative<br>partnerships with people from other<br>countries | Yes | 148<br>(89.2) | 18<br>(10.8) | - | - |
|  | No | 14<br>(66.7) | 7<br>(33.3) | 4.11 (1.40-11.34,<br>p=0.007) | 2.71 (0.81-8.77,<br>p=0.096) |

**Table 3:** Agreement on the rights of local and collaborating scientists, authorship and benefit sharing in collaborative research

| Dependent: Involved in the collection of human biological samples for use in future research | Observation | Yes | No |
| --- | --- | --- | --- |
| Gender | Male | 92<br>(56.8) | 8<br>(32.0) |
|  | Female | 70<br>(43.2) | 17<br>(68.0) |
| Have ever donated biological samples for research purposes | Yes | 41<br>(25.8) | 6<br>(24.0) |
|  | No | 118<br>(74.2) | 19<br>(76.0) |
| Have ever worked in collaborative partnerships with people from other countries | Yes | 148<br>(91.4) | 18<br>(72.0) |
|  | No | 14<br>(8.6) | 7<br>(28.0) |
| Agree Donors should only be able to provide consent to future research on their samples If the donor will be notified if any information relevant to the donor's health is discovered | Disagree | 40<br>(24.8) | 7<br>(28.0) |

| Dependent: Involved in the collection of human biological samples for use<br>in future research | Observation | Yes | No |
| --- | --- | --- | --- |
|  | Agree | 121<br>(75.2) | 18<br>(72.0) |
| There should be no limitation on the advance consent that donors may<br>provide, so long as future research on their samples is approved by an<br>ethics committee | Disagree | 74<br>(46.2) | 17<br>(68.0) |
|  | Agree | 86<br>(53.8) | 8<br>(32.0) |
| Agree that Samples should always be kept in the country where they<br>were collected | Disagree | 107<br>(66.9) | 14<br>(58.3) |
|  | Agree | 53<br>(33.1) | 10<br>(41.7) |
| Agree that Samples should only be transferred when research facilities<br>are unavailable in their country of origin | Disagree | 41<br>(25.6) | 7<br>(28.0) |
|  | Agree | 119<br>(74.4) | 18<br>(72.0) |
| Agree that If samples are removed from their country of origin, a portion<br>must be left behind so that local scientists can use them for their own<br>research, unless special governmental permission is obtained | Disagree | 36<br>(22.5) | 7<br>(28.0) |
|  | Agree | 124<br>(77.5) | 18<br>(72.0) |
| Agree that MTAs should require that foreign collaborating scientists<br>consult local scientists before any new use of the samples in research | Disagree | 13<br>(8.1) | 4<br>(16.0) |
|  | Agree | 147<br>(91.9) | 21<br>(84.0) |
| Agree that MTAs should require that local scientists have some decision-making power regarding the future use of samples | Disagree | 12<br>(7.5) | 4<br>(16.0) |
|  | Agree | 148<br>(92.5) | 21<br>(84.0) |
| Agree that MTAs should require that decisions regarding future use of<br>samples be made jointly by a committee composed of representatives of<br>both the local scientists and the foreign collaborating scientists | Disagree | 26<br>(16.1) | 1 (4.0) |
|  | Agree | 135<br>(83.9) | 24<br>(96.0) |
| Agree that MTAs should require that local scientists have of veto power<br>over any future use of samples by the foreign collaborating scientists | Disagree | 37<br>(23.0) | 7<br>(29.2) |
|  | Agree | 124<br>(77.0) | 17<br>(70.8) |
| Agree that MTAs should require that a local scientist is involved in the<br>protocol development team for any future research on the samples | Disagree | 23<br>(14.3) | 2 (8.0) |
|  | Agree | 138<br>(85.7) | 23<br>(92.0) |
| Agree that MTAs should require that, in exchange for providing the<br>samples, local scientists are credited for authorship on all publications<br>arising from research on the samples | Disagree | 28<br>(17.4) | 9<br>(36.0) |
|  | Agree | 133<br>(82.6) | 16<br>(64.0) |
| Agree that MTAs should require that, in exchange for providing the<br>samples, local scientists are credited for authorship on the first<br>publication arising from research on the samples | Disagree | 76<br>(47.8) | 10<br>(41.7) |
|  | Agree | 83<br>(52.2) | 14<br>(58.3) |
| Agree that MTAs should require that, in exchange for providing the<br>samples, local scientists are credited for authorship only if local scientists<br>provide sufficient intellectual input into the publication | Disagree | 94<br>(58.8) | 10<br>(40.0) |
|  | Agree | 66<br>(41.2) | 15<br>(60.0) |
| Agree that MTAs should require that local scientists be given the<br>opportunity to provide sufficient intellectual input to be credited for<br>authorship on publications arising from research on the samples | Disagree | 19<br>(11.9) | 4<br>(16.0) |
|  | Agree | 141<br>(88.1) | 21<br>(84.0) |
| Agree that MTAs should require that foreign collaborating scientists<br>share royalties from discoveries, patents, and intellectual property that<br>arises from research on the samples with local scientists | Disagree | 19<br>(11.9) | 5<br>(20.0) |
|  | Agree | 140<br>(88.1) | 20<br>(80.0) |
| Agree that MTAs should require that foreign collaborating scientists<br>share royalties from discoveries, patents, and intellectual property that<br>arises from research on the samples with the population or country from<br>which the samples were taken | Disagree | 44<br>(27.5) | 3<br>(12.0) |
|  | Agree | 116<br>(72.5) | 22<br>(88.0) |
| Agree that MTAs should require that the population or country from<br>which the samples were taken is given access to material products, such<br>as pharmaceuticals, that arise from research on the samples | Disagree | 33<br>(21.0) | 1 (4.0) |
|  | Agree | 124<br>(79.0) | 24<br>(96.0) |
| Agree that Local scientists are under pressure to accept unfavorable<br>conditions for the transfer of their sample collections to foreign<br>collaborating scientists with access to more resources | Disagree | 90<br>(56.2) | 12<br>(50.0) |
|  | Agree | 70<br>(43.8) | 12<br>(50.0) |
| Agree that Binding regulations should be in place to ensure that certain<br>protections for local scientists are included in MTAs | Disagree | 8 (5.0) | 3<br>(12.5) |
|  | Agree | 153<br>(95.0) | 21<br>(87.5) |
| If their samples will be anonymous and unlinked to identifying<br>information | Disagree | 38<br>(23.6) | 6<br>(24.0) |
|  | Agree | 123<br>(76.4) | 19<br>(76.0) |
| If donors will have the opportunity to withdraw consent later on | Disagree | 46<br>(28.6) | 6<br>(24.0) |
|  | Agree | 115<br>(71.4) | 19<br>(76.0) |

Table 3 shows respondents’ level of agreement on the rights of local and collaborating scientists in regards to biobanking, authorship, benefit sharing and what should be included in MTAs. A majority of respondents (73.3%) disagreed with always keeping donated samples in the country where they were collected; however, 75.9% agreed that a portion of the samples must be left behind for the benefit of local scientists. An overwhelming majority were in agreement that local scientists should be involved in decisions making regarding the future use of samples, including protocol development, as shown in table 3. They also agreed that local researchers and populations should get a share of the benefits that arise from the use of the samples for research. They offered various views on the authorship rights of local scientists as shown in table 3. They also indicated that there should be binding regulations to protect local scientists in collaborative research.

In table 4, it is important to note that females were more likely than male respondents to be involved in the process of collecting human biological samples for use in future research.

**Table 4:** Comparisons based on reported involvement in sample collection research

| Dependent: Involved in the collection of human biological samples for use in future research | Adjusted Odds ratio | SE | statistic | P-value | Conf. low | Conf. high |
| --- | --- | --- | --- | --- | --- | --- |
| (Intercept) | 0.04 | 1.96 | -1.70 | 0.09 | 0.00 | 0.82 |
| Gender is female | 6.95 | 0.73 | 2.66 | 0.01 | 1.84 | 33.87 |
| Have never donated biological samples for research purposes | 0.60 | 0.70 | -0.73 | 0.47 | 0.15 | 2.52 |
| Have never worked in collaborative partnerships with people from other countries | 16.47 | 0.89 | 3.16 | 0.00 | 3.07 | 106.46 |
| Agree Donors should only be able to provide consent to future research on their samples If the donor will be notified if any information relevant to the donor's health is discovered | 0.33 | 0.72 | -1.54 | 0.12 | 0.08 | 1.38 |
| Agree that Samples should always be kept in the country where they were collected | 3.90 | 0.70 | 1.95 | 0.05 | 1.04 | 16.87 |
| Agree that Samples should only be transferred when research facilities are unavailable in their country of origin | 0.78 | 0.79 | -0.31 | 0.76 | 0.17 | 4.08 |
| Agree that If samples are removed from their country of origin, a portion must be left behind so that local scientists can use them for their own research, unless special governmental permission is obtained | 0.33 | 0.83 | -1.34 | 0.18 | 0.06 | 1.75 |
| Agree that MTAs should require that foreign collaborating scientists consult local scientists before any new use of the samples in research | 0.02 | 1.36 | -2.85 | 0.00 | 0.00 | 0.28 |
| Agree that MTAs should require that local scientists have some decision-making power regarding the future use of samples | 0.15 | 1.14 | -1.65 | 0.10 | 0.01 | 1.36 |
| Agree that MTAs should require that decisions regarding future use of samples be made jointly by a committee composed of representatives of both the local scientists and the foreign collaborating scientists | 29.45 | 1.57 | 2.16 | 0.03 | 2.10 | 1,271.78 |
| Agree that MTAs should require that local scientists have of veto power over any future use of samples by the foreign collaborating scientists | 1.23 | 0.87 | 0.24 | 0.81 | 0.24 | 7.69 |
| Agree that MTAs should require that a local scientist is involved in the protocol development team for any future research on the samples | 63.17 | 1.87 | 2.22 | 0.03 | 2.54 | 3,620.46 |
| Agree that MTAs should require that, in exchange for providing the samples, local scientists are credited for authorship on all publications arising from research on the samples | 0.17 | 0.94 | -1.89 | 0.06 | 0.02 | 1.06 |
| Agree that MTAs should require that, in exchange for providing the samples, local scientists are credited for authorship on the first publication arising from research on the samples | 0.69 | 0.65 | -0.57 | 0.57 | 0.18 | 2.48 |
| Agree that MTAs should require that, in exchange for providing the samples, local scientists are credited for authorship only if local scientists provide sufficient intellectual input into the publication | 1.21 | 0.76 | 0.25 | 0.80 | 0.26 | 5.39 |
| Agree that MTAs should require that local scientists be given the opportunity to provide sufficient intellectual input to be credited for authorship on publications arising from research on the samples | 0.42 | 1.18 | -0.74 | 0.46 | 0.04 | 4.82 |
| Agree that MTAs should require that foreign collaborating scientists share royalties from discoveries, patents, and intellectual property that arises from research on the samples with local scientists | 0.21 | 1.06 | -1.45 | 0.15 | 0.03 | 1.90 |
| Agree that MTAs should require that foreign collaborating scientists share royalties from discoveries, patents, and intellectual property that arises from research on the samples with the population or country from which the samples were taken | 21.09 | 1.35 | 2.26 | 0.02 | 2.20 | 470.50 |
| Agree that MTAs should require that the population or country from which the samples were taken is given access to material products, such as pharmaceuticals, that arise from research on the samples | 12.73 | 1.42 | 1.80 | 0.07 | 1.22 | 442.27 |
| Agree that Local scientists are under pressure to accept unfavorable conditions for the transfer of their sample collections to foreign collaborating scientists with access to more resources | 1.08 | 0.67 | 0.11 | 0.91 | 0.29 | 4.05 |
| Agree that Binding regulations should be in place to ensure that certain protections for local scientists are included in MTAs | 0.14 | 1.45 | -1.37 | 0.17 | 0.01 | 2.23 |

This was significant (Adjusted Odds Ratio=6.95, p value = 0.01). The respondents who were involved in collecting human biological samples for future research were: (a) more likely to never have worked in collaborative partnerships with people from other countries. This was significant (Adjusted Odds Ratio=16.47, p value = 0.002). (b) significantly more likely to agree that samples should be kept in the country where samples were collected

(Adjusted Odds Ratio=3.9, p value = 0.05). (c) Significantly less likely to agree with having MTAs requiring foreign collaborating scientists to consult local scientists before any new use of samples in research (Adjusted Odds Ratio=0.02, p value = 0.004). (d) significantly more likely to agree that MTAs should require that future decisions regarding future use of samples should be jointly made by a committee composed of representatives of both the local scientists and the foreign collaborating scientists (Adjusted Odds Ratio=29.45, p value = 0.03). (e) Significantly more likely to agree that the MTAs should require a local scientist is involved in the protocol development team for any future research on the samples (Adjusted Odds Ratio=63.17, p value = 0.03). and (f) significantly agreed that MTAs should require foreign collaborating scientists to share the royalties from patents and discoveries and intellectual property that arises from the research on the samples with the population or the country from which the samples were taken (Adjusted Odds Ratio=21.09, p value = 0.02).

### Summary of respondents reported experiences with MTA’s

There were 81 responses to the open-ended item asking respondents to “write any comments you might have about your experience with MTA’s.” As shown in figure 2, the word cloud, the five most common words used in the statements on participants experiences were” MTA’s (52 occurrences), Samples (45 occurrences), Scientists (38 occurrences), local (33 occurrences) and transfer (20 occurrences). Overall the responses were more positive than negative (proportion of words with a positive sentiment = 59.04%). The positive sentiments reduced with each unit increase in the age of respondents. This was not significant (Odds ratio=0.98, p value = 0.6). Also, female respondents were less likely to have positive sentiments in their MTA related experiences compared to male respondents. This was not significant (Odds ratio= 0.89, p value = 0.8). In the next paragraphs are summaries of the participants responses are presented under two themes the “MTA process” and “Trust issues in the Process.”

**Figure 2:**
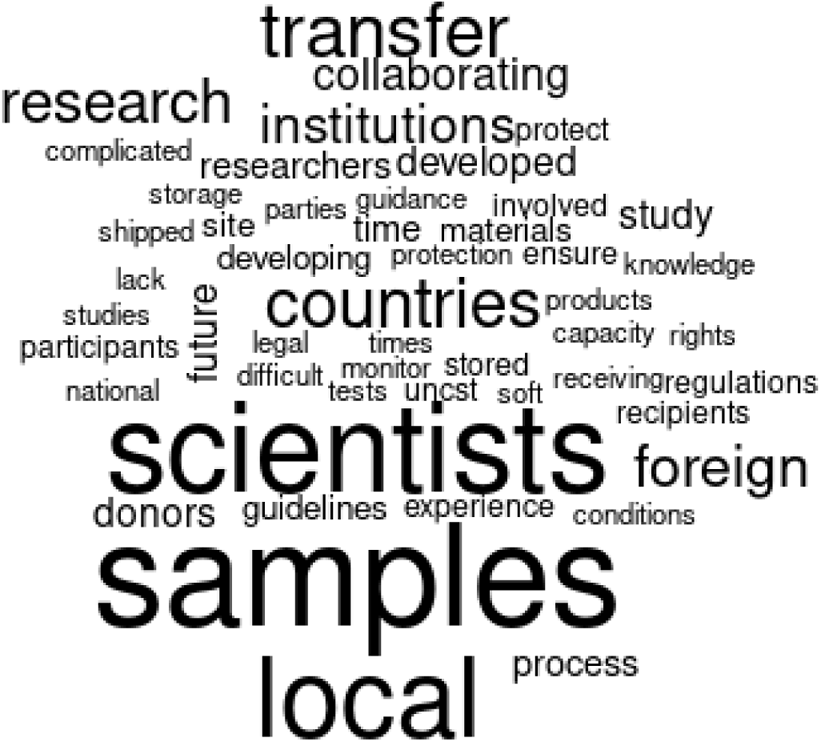
Word Cloud from content analysis of participants responses

## Theme 1: The MTA process

The first theme, the “MTA process,” has suggestions on how to improve the current MTA process as a whole, to include current experiences and potential interventions. There were three codes under this theme:(1) Capacity of the local institutions, (2) Guidelines and legal help and (3) the MTA experience. Each of these codes is described in more detail in the next paragraphs.

### Capacity of local institutions

There were 30 sections of text coded under this code that focused on suggestions for the enhancement of the protection role and/or empowerment of the institutions that either host or regulate local scientists. Training of local scientist by the regulatory agencies or their host institutions was identified as one of the key interventions. It was suggested that this training should include details of what is expected in an MTA with an emphasis on reading the MTA documents in detail, instead of simply signing. Other suggestions included defining the roles of local scientists, especially after transfer of the samples across the border. It was noted that a definition of these post transfer roles is currently missing or unclear. Training would give the local scientists a clearer appreciation of the value of both the MTAs and donated samples. It was noted that the need to export samples can be minimized by increased transfer of knowledge and technology to local institutions to empower them to use or develop innovations. Where samples are eventually shipped the local institutions should be given the opportunity to inspect the facilities, as part of a verification process, where the samples will be stored. In addition to ensuring clear and timely communication about the shipped sample, it was suggested that there should be local independent verification bodies other than the RECs and UNCST to vet the MTA related activities. Some respondents noted that some of the local regulatory institutions are a source of delays that sometimes leads to loss of opportunities. In addition, it was noted that the local RECs seem to have no say in the MTA approval currently. At the host institutional level, the absence of local institutional research agendas greatly impacts the direction of the MTA negotiations resulting in MTA’s that are vague and lacking adequate local institutional protections

*“MTAs are usually written by the foreign collaborating scientists which is most likely to be biased towards their interests. Local scientists/regulatory authorities should be given the opportunity to inspect the facilities where samples are stored.”* Female respondent

*“The language should be very clear and the recipient should be in direct contact with the local research …. and any research on the materials must be communicated. Otherwise capacity building in our local laboratories should be advocated for before any signing of an MTA and whenever possible samples should not” be shipped.”* Female respondent

### Guidelines and legal help

There were 24 sections of text coded under this code that focused on local scientists’ need for guidelines and legal help in the MTA process. Some of the respondents noted that there seemed to be no guidelines for the development of MTA’s at both the local, national or international levels to guide researchers. On the other hand, one male researcher said that he was satisfied with the current guidelines. This suggests that the guideline, if present, may not be easily accessible. Other respondents pointed to specific aspects of the MTA like: the need for them to be site specific as opposed to being general, sometimes as being very complicated for beginners, that they need to be clear, define roles and legal responsibilities and place an emphasis on constant open communication. It was also mentioned that the guidelines should include required pre-requisites like the need to have in place collaborative agreements before MTA negotiation is started. Due to the detailed nature of MTAs, oftentimes they are written in complicated legal language and in some jurisdictions, there may be difference in the process of MTA negotiation for different countries even in Africa; several researchers suggested that having legal expertise involved was a key pre-requisite.

*“There is a general lack of solid national guidelines or policy regarding the appropriate or adequate content of the MTA. Non-exhaustive guidelines in UNCST guideline pertaining to the use of biological samples from Uganda. Inability to monitor usage of transferred samples in case of lack of transparency or dishonesty of receiving scientists.”* Female respondent

### MTA experience

There were 6 sections of text coded under this code that focused on local scientists’ experience of the MTA development process. Respondents described the MTA development process as both tedious and slow. Others described the MTA development process as very rigorous with the potential to become complicated. Some of the complications included disappointments leading to loss of research benefits as a result of change of site after actively participating in the negotiations.

*“I was consulted at the time of developing the protocol as the unit head of a research study site I had made inputs for MTA, unfortunately later the research was taken elsewhere and I did not benefit in any way.”* Female respondent

## Theme 2: Trust issues in the MTA development process

This second theme that was on trust issues in the MTA process, is a summary of the participants concerns about various aspects of the development and eventual use of MTAs. The two (2), codes under this theme are sample use and partnerships. A detailed description for each of these codes is given in the next set of paragraphs.

### Sample Use

There were 24 sections of text coded under this code that focused on local scientists concerns about sample use as part of the MTA development process or later implementation. It was noted that there was no way to monitor the way the samples were used at the foreign sites possibly due to lack of follow up mechanisms or clear communication about the shipped samples back to the local researchers and participants. It was noted that the MTAs do not put restrictions on the extent of both the present and future use or number of tests that will be applied to the samples. It seems that because the extent of future use is not defined, some of the mentioned tests are not included in the protocol thus the need to involve local scientists in all stages of sample use. Respondents wondered how the local scientists and communities benefit from exported HBM. Some felt that this is often not clear in the MTAs and that sometimes this may lead to exploitation. This MTA lack of clarity was said to extend to ownership of materials and products like publications and patents. Respondents also noted that there was no way for research participants to get to know the findings of the studies conducted on the samples they had donated. Some respondents thought that consenting carried a seemingly hidden motive with the focus being on the research that will follow as opposed to the study the participant was taking part in. They indicated that this still applies even when research participants have the option to opt out after non-coercive and adequate participant counselling. This may be misunderstood since the real value of the research is really on the later work as opposed to the current study. Some of the respondents wanted to have transfer of left over samples back to the local scientists for own use though this was left as a suggestion given that the process of transfer is very hectic. Overall the local scientists were not sure or did not trust that the MTA’s were upheld to once samples were shipped.

*“The regulations on MTA are soft, thus the recipients are not compelled to give feedback in future. there are no mechanisms to track whether there is adherence to provisions in the MTA by the collaborating scientists”*. Male respondent

*“In one of our studies, permission to use samples by our collaborators was sought for only once. Later they compromised to do studies and proceeded without our permission. This study on the MTA process is vital!”*. Female respondent

### Collaborative partnerships

There were 16 sections of text coded under this code that focused on local scientists concerns about the collaborative partnerships surrounding the MTA development process or later implementation. Respondents felt that most MTAs tend to favor international collaborators or sponsors and do not encourage technology transfer and local capacity building. As a concern, it was also noted that if the MTAs favor local scientists too much it may lead to local academic laziness through “soft landings.” Overall having access to funding, superior negotiating skills or support and other advantages put pressure on the local scientists. This manifests in various situations even where jointly developed MTAs exist the local scientists usually have no control over the agenda for future research. Respondents felt that there is a real risk that some local scientists may be exploited due to financial inducements more so, as many times, the researchers involved in samples shipment were not directly involved in the MTAs’ development. Some researchers felt unequally yoked as collaborators with the local people not knowing what is going on with the samples even when the foreign collaborators communicate. Most times the drafting is done by the foreign site teams with little local input from the local scientists and since these are very detailed documents enforcement is a challenge. It was suggested that MTA’s should be restricted to teams of scientists with equal interests.

*“Sometimes these MT’s are just completed for formality without paying attention to details for the materials transferred especially when the finding is from the foreign collaborating scientist, because the collaborating scientists often have the financial muscle to fund the study, the local scientist will most certainly accept whatever the collaborating scientist might suggest.”* Male respondent.

## Discussion

Our findings revealed that a vast majority of respondents had ever participated in international collaborative biobanking research. However, less than half had ever been actively involved in negotiating MTAs with collaborating institutions. There were several areas of agreement in regard to the rights of local researchers and institutions in collaborative biobanking research and what details should be included in MTAs. Respondents who were involved in collection of HBM for research were more likely to agree that MTAs should require that decisions regarding the future use of samples be done collaboratively by a committee comprising of representatives of both local and foreign collaborating scientists. They opined that protocol development for any future studies should equally be done collaborative. Respondents further contended that MTAs should require foreign collaborating scientists to equitably share the benefits that arise from the research on the samples with the population or the country from which the samples were taken.

There were two prominent themes that emerged from the qualitative data: a) the MTA process and b) trust issues in the MTA negotiation process. Overall, respondents felt that most MTAs are unfair and tend to favor international collaborators or sponsors, and rarely encourage technology transfer and local capacity building. Some respondents felt that the MTA development process was rigorous however they seemed not to be aware of the guidance provided by the Ugandan national ethics guidelines on the cross-border exchange of human biological specimens. They felt that the role of the REC in MTA development was obscure and that local institutions had limited bargaining power to negotiate MTAs that are favorable to their interests. Several trust issues in the MTA development and implementation process were identified. Respondents did not trust that the provisions of the MTA would be respected and upheld by collaborating scientists once the HBM are shipped. They complained about the lack of transparency and dishonesty of receiving scientists; loss of control and lack of adequate mechanisms to monitor the use of exported samples; sample ownership and intellectual property disputes; exploitation and inequitable sharing of research benefits; and authorship challenges.

There was overwhelming support for collaborative partnership in north-south biobanking research where all MTA negotiations, data/sample access and protocol development for any future studies are accomplished by a team comprising of representatives of both local and foreign scientists. Collaborative engagement and partnership between stakeholders in the Global South, and sponsors and researchers from the Global North is very important for biobanking [25]. Collaborative partnership fosters mutual respect and trust between partners; and it is through true collaborative partnership that power differentials and other disparities can be recognized and addressed [25, 47] This seems to be the desired approach by African researchers, Uganda included, that promotes equitable and meaningful research partnerships that minimize disparities and the possibility of exploitation [22].

The transfer of HBM is governed by contractual MTAs between provider and recipient institutions. The purpose of an MTA is to facilitate the exchange of HBM and associated data between institutions, as well as safeguard the interests of sample donors, researchers, and institutions. An MTA also provides guidance on specific ethical-legal principles that must be adhered to when transferring samples [6]. However, our respondents opined that most MTAs are unfair and tend to favor foreign collaborators, and rarely encourage technology transfer and local capacity building. A greater majority contended that the benefits of research should be fairly and equitably shared with the population or country from which the samples were taken. Currently there is hot debate surrounding this issue of benefit sharing in genomic research and biobanking, particularly in LMICs [22,23,48,49]. Benefit sharing is defined by Schroeder [50], as “the action of giving a portion of advantages/profits derived from the use of human genetic resources to the resource providers to achieve justice in exchange, with a particular emphasis on the clear provision of benefits to those who may lack reasonable access to resulting healthcare products and services without providing unethical inducements” (p. 207), may not necessarily be monetary. Non-monetary benefits may include, but are not limited to, human capacity strengthening through training and skills development; community based social projects and public education; providing feedback to research communities; infrastructure capacity building; and sustained access to funding. Research stakeholders expect equitable and fair sharing of the benefits of research with host communities [51]. However, the insistence of developing countries on the inclusion of provisions for benefit-sharing and ways of equitably handling intellectual property rights in standard MTAs has often been rebuffed by Northern research partners [24]. This state of affairs may partly be attributed to the lack comprehensive ethico-legal frameworks to support implementation of benefit sharing in most African countries [8,52,53]. Over the last 10 years, the H3Africa Consortium has endeavored to set an example of how HBM, data and other resources can be ethically shared. It has expended considerable effort in ensuring that HBM and data collected from the various H3Africa projects are appropriately shared. H3Africa established a Data and Biospecimen Access Committee (DBAC) with a cardinal purpose of reviewing and approving or rejecting all requests from the research community, including commercial entities, for access to biospecimens and associated data generated by H3Africa Consortium. This resonates with the suggestion made by our respondents, regarding the establishment of an all-inclusive committee to oversee the future use of HBM and associated data. The DBAC mediates disputes between sample recipients, donating researchers and biorepositories and also ensures that HBM release is done in accordance with the MTA. Some scholars have also offered frameworks that emphasize the respect of African values and cultures, genuine intellectual participation of African stakeholders, and promotion of relationships basing on respect, fairness, equity and reciprocity [54, 55]. This is consistent with the call for collaborative partnership that was suggested by a greater majority of respondents in the current study.

Several respondents in the current study seemed not to be aware of the guidance provided by the Ugandan national ethics guidelines on the cross-border exchange of human biological specimens. This is rather disturbing because a majority of respondents had a research experienced of more than 10 years. It is imperative that all stakeholders, local researchers/institutions included, clearly understand the conditions under which samples can either be transferred across borders, as prescribed by national and international ethico-legal frameworks. For instance, according to the Ugandan national ethical guidelines, HBM can only be exported after demonstrating that in-country capacity to perform the required investigations does not exist or is inadequate, or for quality control and reference purposes. Further, a few respondents were unsure of the responsibilities of RECs in MTA development. This is not surprising because even the Ugandan national ethics and biobanking guidelines do not explicitly pronounce themselves on the role of RECs in biobanking research. Research ethics committees play a very important role in biobank governance. They are responsible for ethical review of processes related to HBM/ data sharing; and ensuring that there is adequate compliance with national and international standards. They also ensure that the interests of the sample donors and the conditions of the informed consent have been met, as well as to guarantee that there is an ethical commitment of both two parties to the MTA to respect those conditions [56]. Therefore, it is important that all stakeholders clearly understand the importance of negotiating MTA that take cognizance of local interests; and RECs play a key role in enforcing this.

Respondents in this study were wary of the loss of control once samples cross borders due to the lack of effective mechanisms for tracking and monitoring their use. They also did not trust that the provisions of the MTA would be respected and upheld by collaborating scientists once the HBM are shipped. We posit that RECs can ably take on this responsibility if they are empowered and adequately facilitated.

The main strength of our study was the combination of both open- and closed ended questions to explore participants’ perceptions and experiences in cross-border transfer of HBM in collaborative research. Our findings are generalizable because participants were drawn from the national database at UNCST, the national research regulator; and a majority were from research intense institutions. We only included one open ended question pertaining to participants’ experiences with negotiating MTAs, and this we feel was a limitation. We should have included more open-ended questions however, this would have made the survey tool long and increased the risk for none-response.

## Conclusion

Overall, local researchers had a positive attitude towards the export of human biological materials in collaborative research. However, there are several governance and trust issues that may be affecting the implementation of biobanking research. Researchers felt that most MTAs are unfair and tend to favor international collaborators, and rarely encourage technology transfer and local capacity building. Much as this may be true, several researchers seemed not to be conversant with the guidance provided by the Ugandan national ethics guidelines on the cross-border exchange of HBM. The national guidelines for biobanking provide several protections for local scientists and institutions; therefore, the lack of awareness could negatively impact their negotiating power during MTA development. Researchers also contended that the benefits of research should be fairly and equitably shared with the population or country from which the samples were taken.

### Best practices

As our findings have indicated, presence of ethico-legal guidance alone is not enough for fair and successful biobanikng research. Stakeholders, particularly researchers, should read and understand the available national and international guidelines, policies and regulations before negotiating MTAs. They should also involve legal experts in MTA development because of the major challenge of contradictory ethico-legal frameworks across borders that may prevent effective implementation of cross-border biobanking research [57, 58]. There is a need for flexibility and agility in these processes if the interests of local scientists and institutions are to be appropriately considered.

### Implications to policy

Ugandan national ethics and biobanking research are not enshrined in law, are brief and lack detail in several aspects. These guidelines are silent on the role of the REC in MTA development, and this has been abused by some researchers. We recommend that if possible, these guidelines should become regulations so that they are legally binding. Further, we believe RECs should be more actively involved in regulatory oversight and compliance monitoring of biobanking research to ensure that the provisions of the MTA are respected and adhered to. The national research regulators and individual institutions should join forces and devise mechanisms for tracking and monitoring the use of exported HBM and data. Institutions should also develop policies and standard operating procedures for MTA development and negotiation in collaborative research.

### Educational implications

Our findings suggest that several researchers were either unaware of the national guidelines for biobanking research or did not understand them. We recommend training of all local stakeholders in research ethics and best practices for biobanking. The UNCST in concert with research/academic institutions should design an educational campaign on the ethical-legal regulatory framework governing biobanking research in the country. This should preferably be included among the activities for the annual research ethics conference in Uganda.

### Research agenda

Our findings have revealed several gaps in biobank governance in Uganda. They have also highlighted pertinent trust issues in collaborative biobanking research. More research should be conducted to develop a better understanding of the biobank governance in Uganda so as to offer evidence based solutions and also inform policy.

## Data Availability

All data produced are available online at https://doi.org/10.6084/m9.figshare.19352303.v3

## Acknowledgements

We are grateful to our respondents for taking time off their busy schedules to participate in this study. We are equally grateful to Deborah Ainembabazi and Godfrey Bagenda our research assistants and Christine Kusasira Maholo for the data entry.

## Author contributions

ESM: Conceptualization, data curation, formal analysis, funding acquisition, investigation, methodology, project administration, supervision, validation, original draft preparation and review and editing.

IGM: Conceptualization, data curation, formal analysis, investigation, methodology, validation, original draft preparation and review and editing.

All authors read and approved the final version of the manuscript.

